# DeepSeek as the paradigm shift in rare disease diagnosis – the power of a fully automated genetic variant classification system

**DOI:** 10.1101/2025.06.03.25328923

**Authors:** Wei Ma, Grace Fong, Joe Lai, Heidi Wu, Shirley Pik Ying Hue, Jonson Ying, The Hong Kong Genome Project, Annie Tsz Wai Chu, Brian Hon Yin Chung

**Affiliations:** Hong Kong Genome Institute; Hong Kong Special Administrative Region, China; Department of Paediatrics and Adolescent Medicine, School of Clinical Medicine, Li Ka Shing Faculty of Medicine, The University of Hong Kong; Hong Kong Special Administrative Region, China

## Abstract

Large language models (LLMs) have been extensively tested for incorporating into medical applications in recent years, yet their potential in clinical genetics, particularly in diagnosing rare diseases, remains underexplored. Recent advancements in LLMs have improved their reasoning capabilities and transparency, facilitating significant enhancements in clinical workflow designs. The open-sourced DeepSeek model also serves as a cost-effective alternative of top-ranked proprietary reasoning LLMs such as o3-mini-high for genome projects and hospitals that have specific needs in data security. In this study, we developed a framework that fully automates genetic variant classification according to the American College of Medical Genetics and Genomics (ACMG) and the Association for Molecular Pathology (AMP) guidelines and Clinical Genome Resource (ClinGen) recommendations. Two state-of-the art LLMs, DeepkSeek-R1 and o3-mini-high were tested for their performance in variant classification. We demonstrated that through careful prompt engineering and creation of ACMG-rule specific knowledgebases, DeepSeek-R1 outperformed o3-mini-high and achieved high sensitivity and 100% specificity in interpreting ACMG rules that require understanding literature-based evidence. Further testing using 150 variants curated by ClinGen experts, DeepSeek-R1 demonstrated performance on par with human curators. Finally, we showed the framework can be also used for reanalysis using 150 ClinVar variants with conflicting interpretations. Our study provided the first LLM framework capable of fully automated variant classification in the diagnosis of genetic diseases and variant reanalysis.

## INTRODUCTION

Large language models (LLMs) have shown enormous potential in medical applications (*1, 2*). Recently released new generations of LLMs have significantly enhanced reasoning capabilities and are anticipated to have even greater power in processing complex biological/medical information. In diagnosis of rare diseases, whole genome sequencing (WGS) has proven pivotal in ending the diagnostic odyssey of many patients. To manage the huge number of variants identified through WGS, efforts have been made by entities such as the American College of Medical Genetics and Genomics (ACMG), the Association for Molecular Pathology (AMP) and the Clinical Genome Resource (ClinGen) working groups to establish guidelines and recommendations (https://clinicalgenome.org/working-groups/sequence-variant-interpretation/) which now form the most widely adopted evidence-based variant classification system (*3*). These systems require extensive expertise and labor-intensive review of evidence, presenting a significant bottleneck in genetic diagnostics. The Hong Kong Genome Project (HKGP), along with numerous global genetic laboratories, has been addressing this challenge through the development of an automated classification workflow that facilitates human-AI collaboration. In this context, LLMs are invaluable for analyzing large volumes of textual data, summarizing relevant evidence, and suggesting appropriate ACMG rules to geneticists and curators (*4*). Studies have demonstrated that through applying prompting methods such as prompt tuning and retrieval-augmented generation (RAG), LLMs can outperform human experts in certain medical tasks (*1, 5*). However, the integration of LLMs into clinical genetics is not without challenges. Issues such as artificial hallucination and overthinking can impact the accuracy of variant classifications, as these errors may lead to the provision of incorrect evidence and overdiagnosis. Although strategies like chain-of-thought prompting have been effective in reducing false positives in medical applications, some model-specific hallucinations may still persist and remain difficult to eliminate (*6*). Recent developments have seen LLMs such as DeepSeek-R1 enhance transparency in their reasoning processes, which is crucial for refining their application in medical settings (*7, 8*). The open-sourced nature of DeepSeek-R1 also gives it advantage in application for genome projects such as the Hong Kong Genome Project which mandates local data retention for security purposes (*9*). Given that DeepSeek-R1 matches the reasoning capabilities of proprietary models such as OpenAI’s o3-mini-high, it serves as an attractive candidate for application in genetic diagnosis using the ACMG classification system which remains an underexplored topic in medical LLM. Previous studies have begun to explore the potential of LLMs in ACMG classification, yet they often covered only a limited number of ACMG rules and still heavily relied on manual interpretation to reach final decisions (*10, 11*). This highlights a significant opportunity for more comprehensive research to leverage LLMs in automating and enhancing the accuracy of genetic variant classification processes. In this study, we focus on optimizing and evaluating the performance of DeepSeek-R1 and o3-mini-high in summarizing literature evidence and applying this information to variant classification. We have developed a comprehensive framework to fully automate ACMG variant classification and assessed its effectiveness with 150 expert-curated variants from ClinGen and 150 ClinVar variants with conflicting classifications.

## RESULTS

### Establishing a fully automated variant classification framework using LLM

Prior to utilizing LLMs, we have already automated 65% (13/20) ACMG rules that rely on searching publicly accessible databases such as gnomAD and ClinVar and scores from prediction tools like autoPVS1 (for loss-of-function variants), REVEL (for missense variants), and SpliceAI (for splicing variants) within the HKGP (Figure 1A). The remaining 35% (7/20) of the rules, which require summarizing evidence from scientific publications and determining the applicable ACMG rule and its strength, posed challenges for automation. They include evidence for (i) mRNA splicing evidence (PVS1(RNA)), (ii) *de novo* occurrence (PS2/PM6), (iii) experimental evidence (PS3), (vi) prevalence in affecteds (PS4), (v) *in trans* with other pathogenic variants (PM3), (vi) cosegregation (PP1) and (vii) patient phenotype specificity (PP4). To overcome these, we employed LLMs to assist in the literature review process. ACMG rule-specific prompts and knowledgebases for RAG were developed to ensure stable and accurate outputs from LLMs while minimizing hallucinations (Figure 1B). Initial tests with less powerful models, DeepSeek-V3 and o3-mini, revealed significantly lower performance compared to DeepSeek-R1 and o3-mini-high; hence, only the latter models were included in the current study (Data not shown). For prompt engineering, we followed previously published recommendations to construct the initial prompt for each ACMG rule (*12*). We then selected 150 well-curated variants for each of the 6 literature-dependent ACMG rules and 100 curated splicing variants for PVS1(RNA) from the HKGP, comprising approximately 50 rule-positive cases and 100 rule-negative cases (Supplementary Tables 1-7). They were curated by experienced variant curators and reviewed with clinicians to confirm the accuracy of the evidence and the application of ACMG rules. These variants underwent two cycles of analysis with DeepSeek-R1 and o3-mini-high. The initial cycle involved prompt optimization based on the reasoning processes (provided only by DeepSeek-R1) and outcomes of the LLMs.

**Figure 1:**
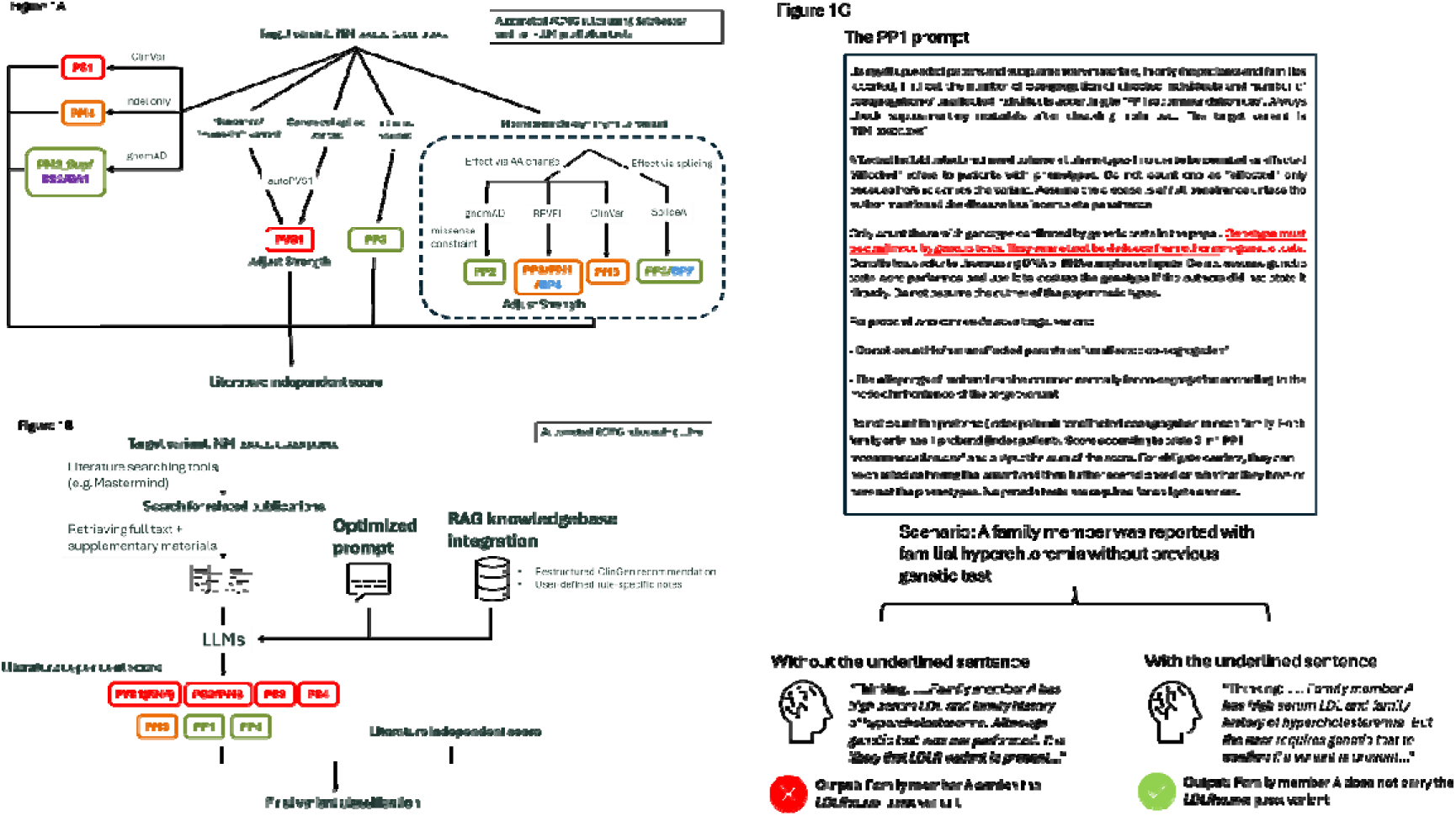
Schematic diagram of overall design of the fully automated ACMG framework. (A) Schematic diagram explaining how literature evidence-independent ACMG rules are applied in the current framework. gnomAD is used to retrieve variant allele frequencies in different populations. ClinVar is used to search for reported patients with the target variant. autoPVS1 is a prediction tool that classifies the effect of different loss of function variants according to the ACMG guideline. REVEL is a machine learning tool that predicts the degree of functional damaging effect of missense variants. SpliceAI is used to predict the potential of a variant in affecting mRNA splicing. (B) Schematic diagram explaining how literature evidence-dependent ACMG rules are applied in the current framework. (C) The PP1 prompt. PP1 rule is applied for evidence of variant co-segregating with disease. LLM is asked to count the number of affected and unaffected segregations based on genotype and phenotype of family members. Sentences to suppress hallucinations in patient genotype are underlined and highlighted in red. A case of determining the genotype of a family member is provided as an example. Without the sentence, LLM may infer family member A having a LDLR variant using results from serum LDL level and family history.

Notably, many artificial hallucinations we encountered remained challenging to address without understanding their underlying causes (*13*). For example, DeepSeek-R1 occasionally incorrectly suggested that a patient carried a specific variant, even though no genetic test had been performed. Upon examining the model’s reasoning steps, we discovered that DeepSeek-R1 was inferring a patient’s genotype based on a high correlation between laboratory test results and the characteristics of a rare disease. This issue was resolved by refining the prompt to explicitly state that “genetic test must be performed in order to determine the genotype of the patient” (Figure 1C). However, prompt tuning alone was not always sufficient to overcome these challenges and RAG knowledgebases were needed to further improve the performance. For rules that could not be optimized solely through prompt engineering, particularly those requiring an assessment of the consistency and specificity of patient phenotypes (such as PS2/PM6, PP1 and PP4), we constructed knowledgebases consisting of restructured ACMG recommendations and manually curated gene-disease/phenotype associations for the RAG pipelines to enhance LLM performance (Supplementary Tables 8-10) (*14, 15*). With contributions from ClinGen, some of the genes have already had expert-curated criteria over the general ACMG guideline. We have compiled a comprehensive list of all genes with ClinGen expert panel recommendations and genes with rule-positive variants identified in the HKGP into our RAG knowledgebases. This list can be updated as new rule-positive variants are identified. Another complex scenario involved determining the application strength of PS4 based on the number of probands reported in the literature carrying the same variant. After reviewing thresholds proposed by all currently available ClinGen expert panels, which were based on disease prevalence and phenotype specificity, we found no consensus (Supplementary Table 11). In our model, which prioritizes sensitivity, we applied a universal threshold (Supporting≥ 1, Moderate≥ 2, Strong≥ 4). For scenarios where maximizing specificity is crucial, thresholds can be adjusted to (Supporting ≥ 2, Moderate≥4, Strong≥16). The accuracy of LLMs was significantly improved by using these user-defined RAG knowledgebases.

After optimization, the performance of DeepSeek-R1 and o3-mini-high was reassessed using the finalized prompts and RAG pipelines on their ability to summarize literature evidence for ACMG classification. In our assessment of the seven literature-dependent ACMG rules, DeepSeek-R1 consistently outperformed o3-mini-high in sensitivity for all tested rules, while both models demonstrated perfect specificity and high concordance with expert interpretations (Table 1). Since only DeepSeek-R1 provided transparency in its reasoning process, the optimization of prompts was based on its thinking process and the final outputs from both LLMs. This underscores the importance of future reasoning LLMs to disclose their reasoning process for better incorporation in medical applications. Notably we have achieved high accuracy only after one round of optimization. Theoretically, the performance of LLMs can be further improved via more rounds of refinement. Remarkably, DeepSeek-R1 achieved 100% sensitivity for evidence related to mRNA splicing (PVS1(RNA)) and patient phenotype specificity (PP4) [o3-mini-high: 97.3% and 93.3%, respectively]. This is because studies related to these evidence often discussed the effect of the variant and the phenotypes associated with the patients in detail. LLMs can therefore clearly catch the key features and apply such information to the corresponding ACMG rules effectively. On the other hand, both LLMs exhibited relatively weaker sensitivity in interpreting evidence for *in trans* occurrence (PM3) [DeepSeek-R1: 88.2%; o3-mini-high: 72.4%] and co-segregation (PP1) [DeepSeek-R1: 73.8%; o3-mini-high: 60.7%]. For PM3, a common issue was the frequent miscounting of the number of patients reported in studies. Notably, many of the studies only mentioned individual patient genotypes once in one of the tables which increased the likelihood of this information being lost or incorrectly extracted during the tokenization of input PDF files. Additionally, both models struggled with patient information presented in complex tables and pedigrees, which require visual interpretation. These formats pose challenges that neither DeepSeek-R1 nor o3-mini-high can handle currently.

**Table 1.** Evaluation of the performance of DeepSeek and o3-mini-high in literature-dependent ACMG rules.

|  | Deepseek-R1 |  |  | o3-mini-high |  |  |
| --- | --- | --- | --- | --- | --- | --- |
|  | Sensitivity | Specificity | Overall accuracy | Sensitivity | Specificity | Overall accuracy |
| PVS1 (RNA)<br>Splicing evidence | 1.000 (37/37) | 1.000 (63/63) | 1.000 (100/100) | 0.973 (36/37) | 1.000 (63/63) | 0.990 (99/100) |
| PS2/PM6<br>*de novo* occurrence | 0.976 (40/41) | 1.000 (109/109) | 0.993 (149/150) | 0.902 (37/41) | 1.000 (109/109) | 0.973 (146/150) |
| PS3<br>Experimental<br>evidence | 0.984 (63/64) | 1.000 (86/86) | 0.993 (149/150) | 0.797 (51/64) | 1.000 (86/86) | 0.932 (137/150) |
| PS4<br>Prevalence in<br>affecteds | 0.940 (94/100) | 1.000 (50/50) | 0.960 (144/150) | 0.910 (91/100) | 1.000 (50/50) | 0.940 (141/150) |
| PM3<br>In trans with other<br>pathogenic variants | 0.882 (67/76) | 1.000 (74/74) | 0.940 (141/150) | 0.724 (55/76) | 1.000 (74/74) | 0.860 (129/150) |
| PP1<br>Co-segregation | 0.738 (45/61) | 1.000 (89/89) | 0.893 (134/150) | 0.607 (37/61) | 1.000 (89/89) | 0.840 (126/150) |
| PP4<br>Patient phenotype<br>specificity | 1.000 (45/45) | 1.000 (95/95) | 1.000 (150/150) | 0.933 (42/45) | 0.968 (92/95) | 0.893 (134/150) |

### Validation of the automated framework using ClinGen-curated variants

After the initial optimization and evaluation of the LLM framework which showed remarkable performance, validation was conducted with 150 ClinVar 3-star variants curated and endorsed by ClinGen expert panels. The interpretation of literature evidence supporting these classifications is detailed in the ClinGen Evidence Repository. Since they were curated using gene-specific recommendations published by ClinGen expert panels which are modified from the original ACMG guidelines, we reassessed the listed evidence and aligned the ACMG rules applied with the original guidelines to ensure a fair comparison of the results generated by LLMs. The variant NM_001369369.1(FOXN1):c.880G>A (p.Val294Ile) will be used as an example to illustrate the power of our workflow (Figure 2A). The literature-independent component of our pipeline first identified that another missense variant, c.880G>C (p.Val294Leu), affecting the same amino acid codon, was reported as likely pathogenic on ClinVar which suggested this could be a mutational hotspot. Next, the pipeline determined that the c.880G>A variant had a GroupMax filtering allele frequency of 6.800e-7 in the gnomAD v4.1.0 database, indicating that it is extremely rare in control populations. Additionally, the variant was scored 0.731 by the ClinGen-calibrated prediction tool REVEL, classifying it as likely functionally deleterious (*16*). Without the use of LLMs, classification of this variant would have stopped at variant of uncertain significance (VUS), requiring manual interpretation of literature-derived evidence to upgrade it to likely pathogenic. With the integration of LLMs, our pipeline automatically analyzed two relevant publications, providing additional evidence to support the pathogenicity of the c.880G>A variant. First, a study assay demonstrated that the variant resulted in only 18% of the normal activity compared to the wildtype protein using luciferase reporter assay (*17*). It was also reported in a homozygous patient diagnosed with severe combined immunodeficiency (SCID) of the T-B+NK+ subtype. This patient also exhibited specific phenotypes, including alopecia totalis and nail dystrophy. Sequencing of an 18-gene panel for SCID-related genes revealed no other pathogenic variants, strongly implicating c.880G>A as the causative variant (*18*). Furthermore, the variant co-segregated with the disease in a sibling from the same family, who was also homozygous for the variant. By combining these findings from the literature, the NM_001369369.1(FOXN1):c.880G>A (p.Val294Ile) was automatically upgraded to likely pathogenic using our pipeline, without requiring manual interpretation. This demonstrates the power of integrating LLMs into variant classification workflows to streamline and enhance the interpretation of evidence.

**Figure 2:**
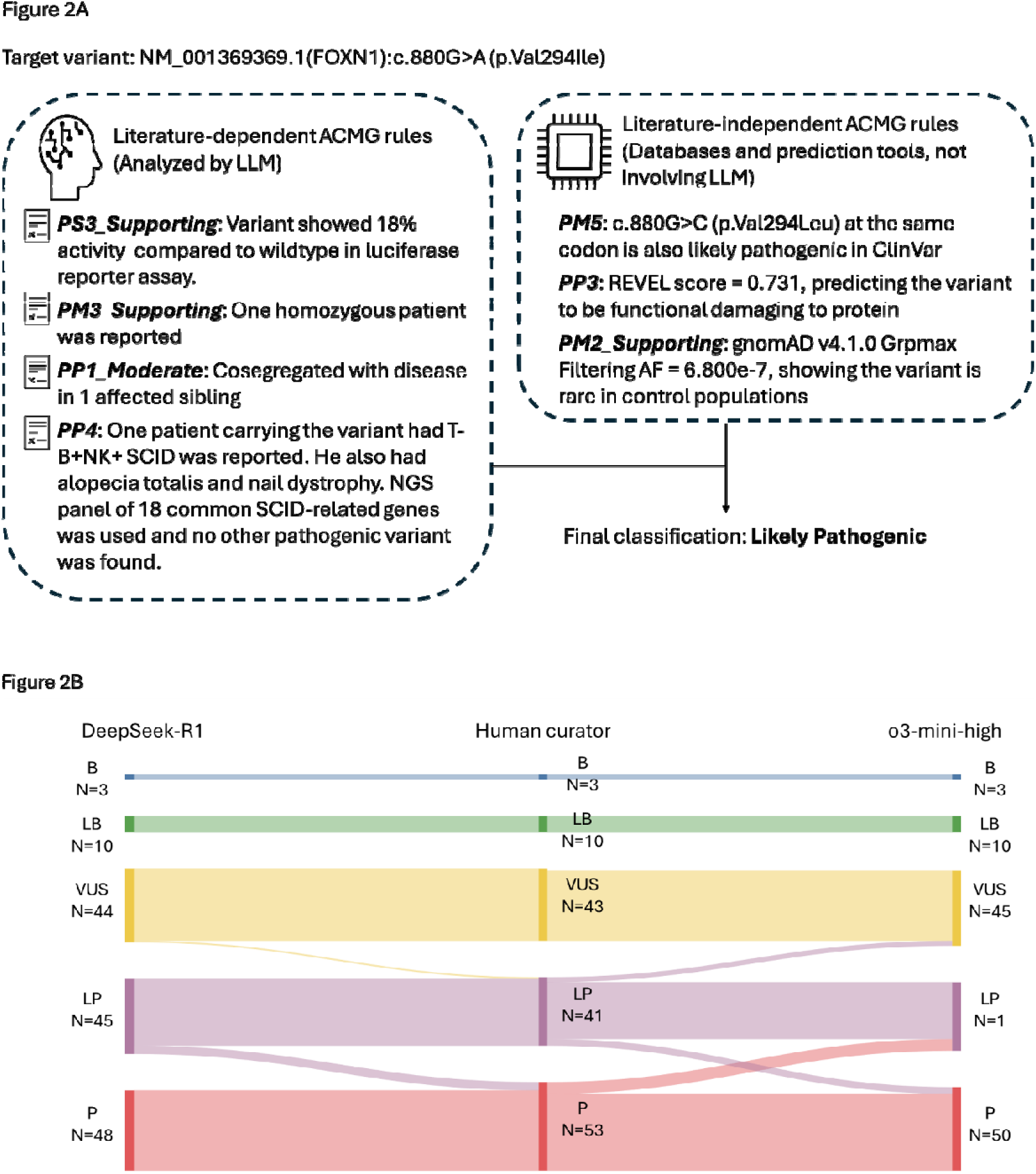
The automated framework in classifying ClinGen variants and reanalysis of ClinVar conflicting variants. (A) The ClinGen curated NM_001369369.1(FOXN1):c.880G>A (p.Val294Ile) variant is used as an example to show the complete analysis using the automated framework. Four different publications were identified and processed by LLM in the literature-dependent arm and resulted in the application of PS3_Supporting, PM3_Supporting, PP1_Moderate and PP4 rules. In the literature-independent arm, ClinVar, gnomAD databases and the prediction tool REVEL were used to apply PM5, PM2_Supporting and PP3 respectively. A final classification of likely pathogenic was achieved after combining all the applied rules. Abbreviation: SCID, severe combined immunodeficiency; Grpmax Filtering AF, GroupMax filtering allele frequency (B) Sankey diagram showing the change in variant classification of 150 ClinGen variants by: DeepSeek-R1, human curator or o3-mini-high. The numbers represent the number of variants in each subgroup. P: pathogenic, LP: likely pathogenic, VUS: variant of uncertain significance, LB: likely benign, B: benign

Overall, DeepSeek-R1 demonstrated superior performance (96.0%, 144/150) in the final variant classification compared to o3-mini-high (91.3%, 137/150) likely due to only reasoning process from DeepSeek-R1 was available for prompt optimization (Figure 2B, Table 2, Supplementary Table 12). DeepSeek-R1 has applied identical ACMG rules as human curators in 84.0% (127/150) [o3-mini-high: 66.7%, 100/150] of cases and wrongly assigned one rule in 15.3% (23/150) [o3-mini-high: 30.0%, 45/150] and two rules in 0.7% (1/150) [o3-mini-high: 2.7%, 4/150] of cases. However, this only led to misclassification of six cases, including five cases from pathogenic to likely pathogenic and one case from likely pathogenic to VUS by DeepSeek-R1. In five cases of misclassification, DeepSeek-R1 failed to count probands carrying the target variant due to the same reasons described in the previous section. Interestingly, in the remaining case of misclassification, NM_005629.4(SLC6A8):c.338G>A (p.Gly113Asp), both ClinGen experts and our curators agreed that a study by van de Kamp et al. demonstrated decreased function of the transporter carrying the missense variant (*19*). However, the experimental result was reported only as a visual figure within the publication. While this conclusion could still be deduced by combining information from the main text and supplementary tables, both DeepSeek-R1 and o3-mini-high failed to interpret this evidence and rejected the use of PS3_Supporting. This suggested that LLMs are still slightly inferior to humans in the most complicated reasoning tasks. However, this could be potentially solved by improvement of LLMs in the future. Since pathogenic variants are often treated similarly as likely pathogenic variants in clinical settings, DeepSeek-R1 achieved a concordance rate of 99.3% (149/150) in genetic diagnosis. In comparison, o3-mini-high misclassified 3 cases from likely pathogenic to VUS which resulted in a concordance rate of 98.0% (147/150). This demonstrated that despite some discrepancies in rule application by LLMs compared to human curators, these differences were generally not significant enough to impact the final diagnosis. Previous studies have shown that human curators across different accredited laboratories exhibit discordance that could affect diagnosis in 11% of tested variants, compared to only 1% in our DeepSeek-R1 framework (*20*).

**Table 2.** Performance of the fully automated ACMG classification framework on 150 ClinGen curated variants.

|  | ACMG rules applied (compared to human curator)* |  |  |  | Final classification (compared to human curator) |  |  |
| --- | --- | --- | --- | --- | --- | --- | --- |
|  | Identical | 1 wrong | 2 wrong | 3 wrong | Identical | Non-identical<br>(Does not affect<br>diagnosis)** | Non-identical<br>(Affect diagnosis)** |
| DeepSeek-R1 | 0.840<br>(127/150) | 0.153<br>(23/150) | 0.007<br>(1/150) | 0.000<br>(0/150) | 0.960 (144/150) | 0.033 (5/150) | 0.007 (1/150) |
| o3-mini-high | 0.667<br>(100/150) | 0.300<br>(45/150) | 0.027<br>(4/150) | 0.007<br>(1/150) | 0.913 (137/150) | 0.067 (10/150) | 0.020 (3/150) |

*\*Rules applied are considered identical only if they have both the same strength and same raw scores as applied by human curators*
*\*\*Final classification of pathogenic and likely pathogenic are considered the same in diagnosis*

### Applying the automated framework for reanalysis of genomic data

We evaluated the potential of applying this fully automated variant classification framework for reanalysis of genome sequencing data. Unresolved ClinVar variants including 93 variants with conflicting classifications and 57 VUSs were selected and re-analyzed by DeepSeek-R1or o3-mini-high, followed by validation by our human curators. Among these variants, DeepSeek-R1 resolved the classification in 70.0% (classification changed to P/LP/LB, 105/150) [o3-mini-high: 66.7%, 100/150] cases, and demonstrated strong concordance with the results of human curators (98.7%, 148/150) [o3-mini-high: 94.7%, 142/150] (Figure 3, Table 3, Supplementary Table 13). Consistent with prior results from the validation section using ClinGen variants, DeepSeek-R1 exhibited errors in the calculation of one ACMG rule in nine cases, resulting in the application of incorrect strength in five cases. In comparison, o3-mini-high demonstrated relatively inferior performance, with errors in the calculation of one ACMG rule in 29 cases and two ACMG rules in one case, leading to incorrect strength application in 17 cases. Again, most errors were attributed to miscounting the number of reported patients carrying the target variant, rather than incorrect data interpretation. LLMs were particularly sensitive in reclassifying variants with conflicting classifications, likely owing to the fact that these variants have accumulated evidence from literature. DeepSeek-R1 correctly upgraded 75.3% (70/93) of those variants to pathogenic/likely pathogenic compared to 69.9% (65/93) by o3-mini-high. DeepSeek-R1 only misclassified one conflicting ClinVar variant, *HIF1A* NM_000545.8:c.467C>T (p.Thr156Met), from VUS to likely pathogenic due to misinterpretation of functional data from a prior study. Although the study reported that the variant caused reduced DNA binding, the authors eventually classified it as neutral (*21*). Such conflicting results within a single study remain challenging to interpret, even for human curators. For ClinVar VUSs, which generally have less supporting evidence compared to variants with conflicting classifications, DeepSeek-R1 also managed to upgrade 50.9% (29/57) to pathogenic/likely pathogenic [o3-mini-high: 45.6%, 26/57]. This not only confirms the efficacy of our framework for primary analysis but also highlights its utility in the reanalysis of variants, demonstrating its broader applicability in genetic diagnostics.

**Figure 3:**
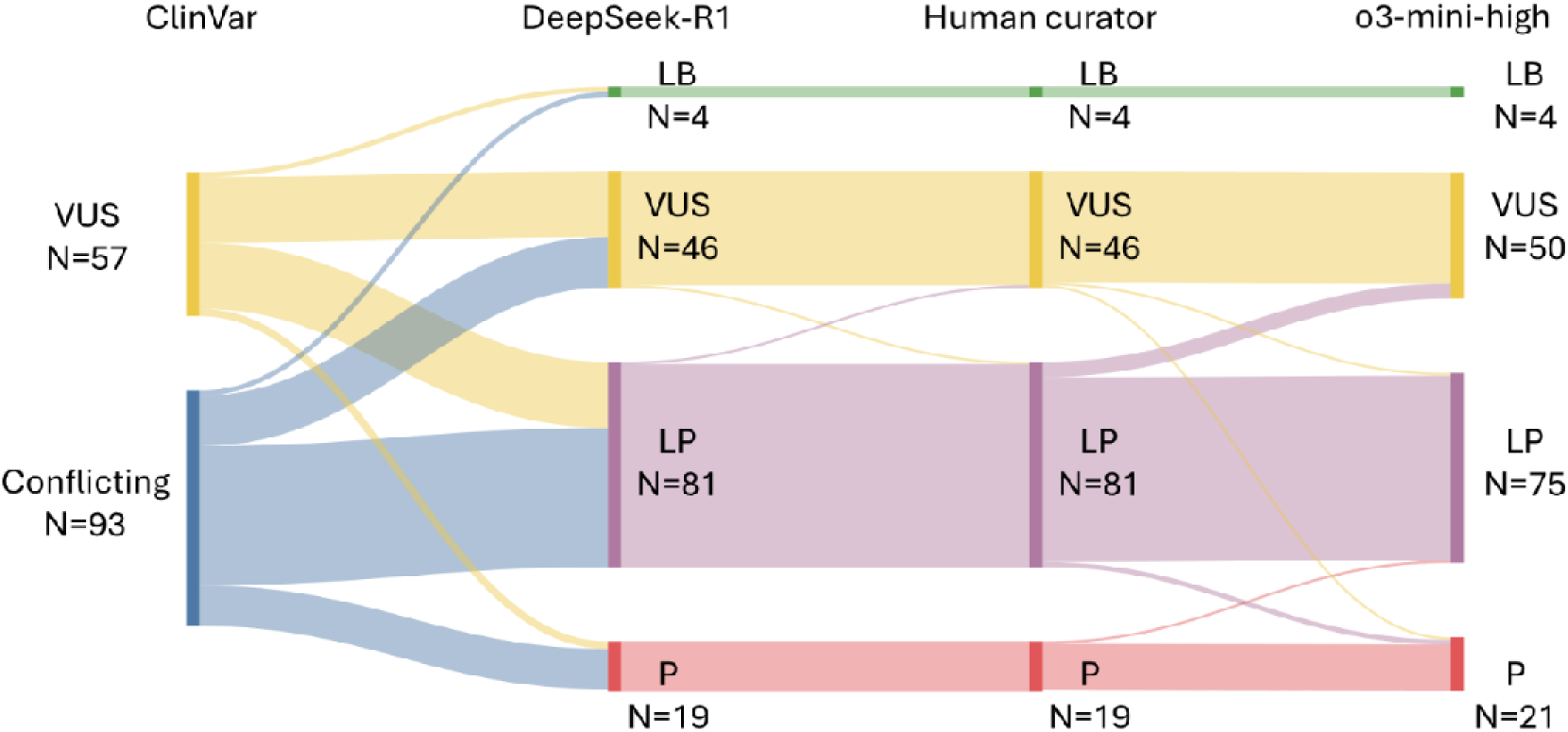
Sankey diagram showing the change in variant classification of 57 VUS and 93 conflicting ClinVar variants by reanalysis done by: DeepSeek-R1, human curator or o3-mini-high. The numbers represent the number of variants in each subgroup. P: pathogenic, LP: likely pathogenic, VUS: variant of uncertain significance, LB: likely benign

**Table 3.**
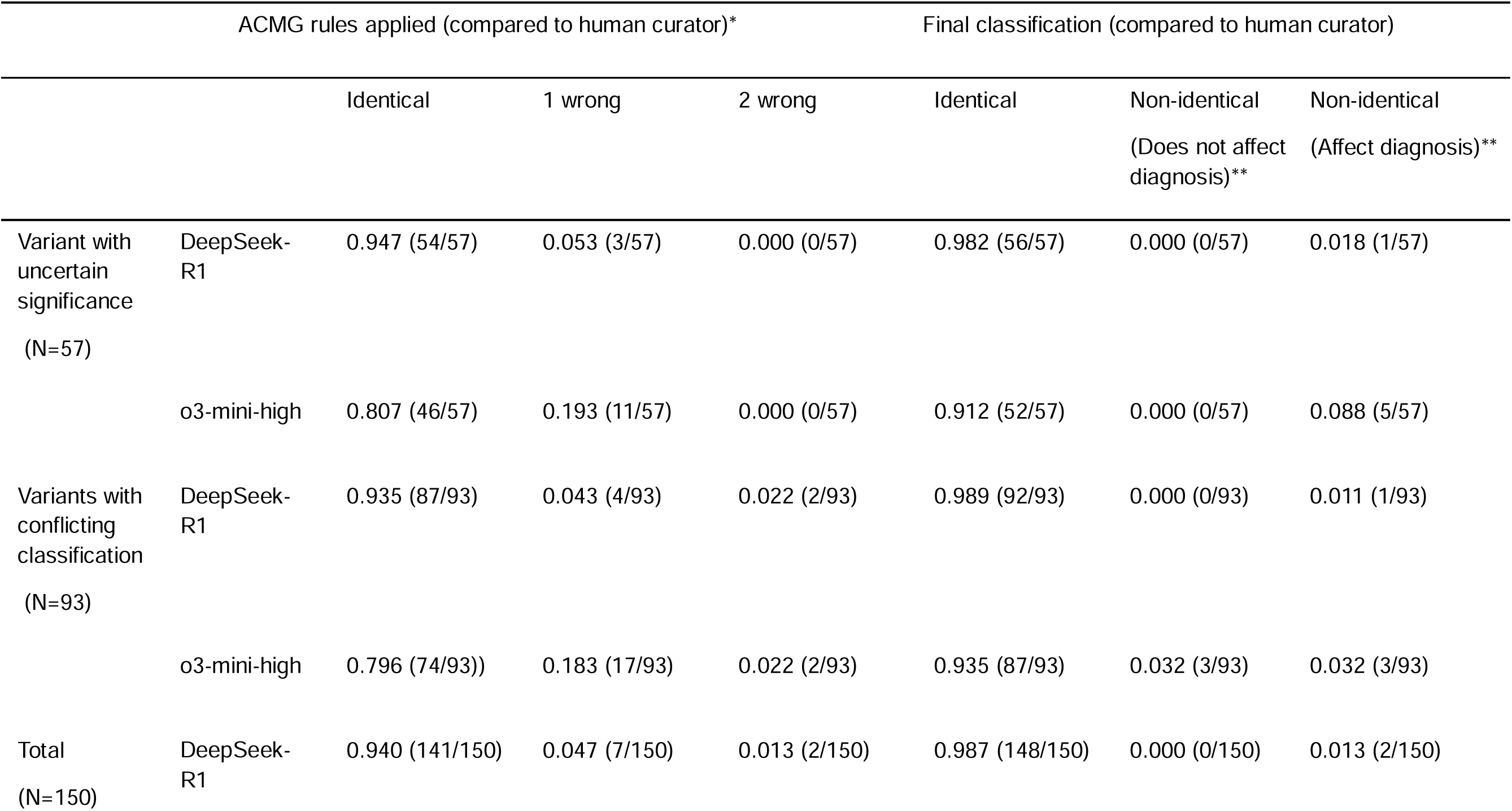
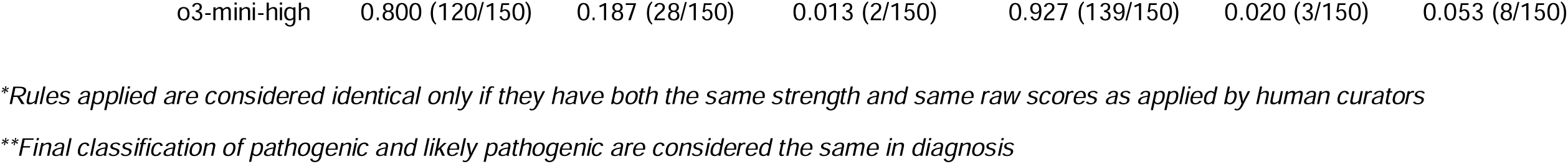
Performance of the fully automated ACMG classification framework on reanalysis of 150 ClinVar variants.

## Discussion

Patients with rare genetic disorders often face prolonged diagnostic odysseys, which come with significant personal and societal challenges. Advances in DNA sequencing technologies have greatly improved throughput and reduced the time required to generate sequencing results. However, identifying and classifying disease-causing variants remain labor-intensive tasks. Efforts have been made to standardize variant classification workflows to shorten analysis time, with the ACMG and ClinGen playing pivotal roles in translating complex evidence-based systems of variant pathogenicity into a Bayesian-based scoring system that is easier to apply. While some ACMG criteria can be automated using databases like gnomAD and ClinVar, as well as bioinformatic tools such as REVEL and spliceAI, many criteria still require manual interpretation of evidence from the literature (*22–24*). This reliance on manual curation slows the diagnostic process, limiting the timeliness of genetic diagnoses for patients. Previous studies have primarily tackled this issue by automating phenotype extraction from electronic health records (EHRs) as evidence for disease specificity (*25–27*). However, this approach risks overestimating phenotypic evidence, particularly for diseases like neurodevelopmental disorders, where patient phenotypes are often nonspecific and with high genetic heterogeneity. Other studies have used experimental results to upgrade variant pathogenicity. Nevertheless, the experiments that can truly represent the mechanism of pathogenicity often require discussion among experts before a consensus could be made. There is again risk of overestimating evidence from a single study to upgrade the variant. Furthermore, critical evidence types such as *de novo* occurrence, *in trans* occurrence with other pathogenic variants for recessive diseases, prevalence in affecteds for dominant diseases, and co-segregation still rely heavily on literature. Without literature-derived evidence, many variants remain classified as VUS.

In this study, we describe a fully automated framework for variant classification that integrates LLMs to handle both literature-independent and literature-dependent evidence. The LLMs, particularly DeepSeek-R1, demonstrated high accuracy (89.3%–100%) in determining the application of seven literature-dependent ACMG rules. The automated system achieved a 99.3% concordance rate with human experts in the final diagnosis of 150 ClinGen-curated variants and a 98.7% concordance rate in resolving ClinVar variants with conflicting classifications or VUS. By optimizing prompts and constructing RAG knowledgebases, the framework effectively prevented hallucinations. Importantly, the LLMs rarely misapplied ACMG rules in the absence of supporting evidence, which limited the risk of overestimating pathogenicity. In the current cohort of 150 ClinGen and 150 ClinVar variants, the framework almost only underestimated pathogenicity, with only one case of overdiagnosis. The use of LLMs significantly streamlined the analysis process. LLMs can finish summarizing the literature evidence in minutes. In comparison, curators in the HKGP typically spent around four hours curating patients with WGS positive findings, with the majority of this time dedicated to reviewing literature. The use of LLMs can significantly enhance analytical efficiency which is critical for certain clinical settings such as rapid molecular diagnosis for critically ill patients and high-throughput clinical applications (*26, 28, 29*). Additionally, an automated framework is essential for the effective periodic reanalysis of unsolved genomic cases, an area that remains a significant challenge in the field (*30–32*). Reanalyzing such cases manually is again labor-intensive, as it often requires curators to review newly published literature. With the expansion of genome projects globally and the adoption of WGS and multiomics approaches, hundreds of new gene-disease associations and thousands of new disease-causing variants are reported annually (*33–35*). Current reanalysis efforts achieve an overall diagnostic yield increase of around 10% without incorporating literature-based evidence (*36*). We believe our automated framework can not only enhance diagnostic yield further but also reduce the costs associated with periodic reanalysis by minimizing the need for human intervention.

Our framework was developed based on the original ACMG guidelines, while incorporating all general recommendations released by ClinGen. Although gene-specific guidelines from ClinGen expert panels were not fully integrated at this moment, our framework offers the flexibility to incorporate them. These can be seamlessly done through simple prompt tuning and the creation of corresponding RAG knowledgebases, further enhancing the adaptability and precision of our LLM-based variant classification system. This adaptability also ensures that the system remains compatible with future updates, including the forthcoming ACMG version 4.0 guidelines, which propose a more structured decision workflow for variant classification. Importantly, our framework can be easily extended to other variant classification systems, such as the ACGS and the ABC systems (*37, 38*). It should be noted that our current framework was designed to handle single nucleotide variants (SNVs) and short insertions/deletions (indels) in Mendelian diseases with moderate to high penetrance. Further development is needed to expand its capabilities to include copy number variants, structural variants, somatic variants, and low-penetrance risk alleles (*39–41*).

Despite its strengths, the automated framework has several limitations. While we have addressed many technical challenges during the optimization phase, some intrinsic limitations of LLMs remain. For instance, LLM accuracy significantly decreases when multiple decisions are required from a single prompt or when the size of the input files are too large. Segmenting tasks into multiple prompts could potentially mitigate this issue, but the exact threshold that triggers performance degradation is not yet clear (*42*). Another limitation is the inability of current LLMs to interpret information from graphical figures and complexly structured tables (*43*). During the optimization of interpreting co-segregation data (PP1), some publications presented family segregation results only in pedigree figures without textual descriptions. Both DeepSeek-R1 and o3-mini-high failed to correctly extract the segregation data. The most recently released LLM, Grok3, was tested and also failed the task (Data not shown). To potentially overcome these challenges, technologies such as optical character recognition (OCR)-GOT model and large vision language models have been suggested (*44, 45*). These advances hold promise for enhancing the performance of LLMs in interpreting complex visual data in future applications. Lastly, at present, our framework processes only evidence from the literature, without integrating patient phenotypes or test results from clinical laboratories stored in the EHR. Most sensitive patient data cannot be uploaded to public servers for LLM analysis. However, the open-source nature of DeepSeek-R1 allows for local deployment in secure environments. This enables us to incorporate AI-based auto-summarization tools of EHR into our automated variant classification framework to further enhance its diagnostic capabilities (*46, 47*).

In summary, we presented here a framework that enables fully automated and accurate variant classification for diagnosis of rare diseases and reanalysis of variants according to the ACMG guidelines and ClinGen recommendations using DeepSeek or LLMs with equivalent reasoning capabilities. It can be easily revised to fit gene-specific rules and new recommendations in the future. Moreover, as newer LLMs increase the transparency in disclosing their reasoning process, model-specific optimization can be readily applied to improve the genetic diagnosis of patients with rare diseases.

## MATERIALS AND METHODS

### Study design

This study was designed to develop and evaluate a framework for fully automated genetic variant classification following the original ACMG/AMP guidelines via the application of LLMs. Without utilizing LLMs, 13 literature-independent AMCG/AMP rules have already been automated through searching publicly available databases and applying various bioinformatic prediction tools. Two state-of-art LLMs, DeepSeek-R1 and OpenAI’s o3-mini-high were optimized to summarize findings in literature to determine the strength of seven literature-dependent ACMG rules that should be applied. Around 50 rule-positive and 100 rule-negative between 1 July 2021 and 31 December 2024 were selected from the HKGP (except for PVS1(RNA), where only 37 rule-positive and 63 rule-negative splicing variants could be identified). They were used for prompt engineering and construction of RAG knowledgebases to enhance and evaluate the performance of the LLMs. The optimized framework was then validated using 150 variants selected from ClinVar 3-star variants which are all curated and endorsed by ClinGen expert panels. The framework was further tested for its capability in reanalysis by applying it to resolve 150 VUSs and variants with conflicting classifications in ClinVar. All publications were analyzed by the LLMs with a minimum of two replicates to ensure data reliability.

### Automation of literature-independent AMCG/AMP rules

The ClinVar database was searched for evidence regarding (i) established pathogenic variants causing the same amino acid change (PS1) and (ii) different pathogenic missense variants affecting the same codon (PM5). The gnomAD database was searched for minor allele frequency (MAF). BA1 and BS1 were applied if the variant has a MAF exceeding 5% and 1% respectively. PM2_supporting was applied if the variant has a MAF≤0.5%. For missense variants, PP2 was applied if the affected gene shows intolerance to missense variation (having a constraint Z score ≥3.09 in gnomAD v2). The bioinformatic tool REVEL was used to predict the effect of missense variants using the following calibrated thresholds: ≤ 0.003, BP4_VeryStrong; (0.003-0.016], BP4_Strong; (0.016-0.183], BP4_Moderate; (0.183-0.290], BP4; [0.644, 0.73), PP3; [0.773-0.932), PP3_Moderate; ≥0.932, PP3_Strong (*16*). SpliceAI was used to predict whether the variant potentially disrupt splicing. PP3 was applied if the variant scored ≥0.2 for both intronic and exonic variants (*48*). Additionally, for exonic synonymous variants, BP7 was applied if it has a SpliceAI score <0.2. PM4 was automatically applied for all variants that result in in-frame changes in protein length. The prediction tool autoPVS1 was used to determine the strength of PVS1 that should be applied for all loss-of-function variants (*49, 50*).

### Optimization and evaluation of LLMs for literature-dependent ACMG rules

Prompts were first designed based on ACMG guidelines and ClinGen recommendations with modification to fit LLMs. The prompts were then optimized using the 150 selected HKGP variants (100 HKGP variants for PVS1(RNA)). The selected variants were all curated by 2 experienced curators in HKGI and the final classification has been endorsed by referring clinicians. Rule-positive variants refer to variants in which the ACMG rule of interest was applied and vice versa. For rule-positive variants, the supporting publications and all of their supplementary materials were all submitted together with the prompt to the LLMs. Reformatting excel files into csv files and word files into text files (txt) were performed as the former ones are not recognized by both LLMs. For rule-negative variants, we searched relevant publications that mentioned the target variant but do not support the application of the rule of interest using the Mastermind search engine (Genomenon). The prompts were optimized according to the reasoning process provided by DeepSeek-R1 and the final output from both DeepSeek-R1 and o3-mini-high. The prompt optimization focused mainly on providing general guidance on handling evidence in the literature to the LLMs instead of providing publication-specific interpretation directions to avoid overfitting. Three knowledgebases for PS2/PM6, PP1 and PP4 were constructed by summarizing both recommendations from ClinGen expert panels and our internal curation experience to guide the LLMs in making the final decision (*51, 52*). The final prompts and the application workflow are described in detail in Supplementary Figures 1-7 and Supplementary Methods. The finalized prompts were reassessed in the selected HKGP curated variants.

### Validation of the optimized framework

To evaluate our proposed framework in real-world data, 150 ClinGen curated variants with diverse classification (10 benign, 15 likely benign, 25 VUS, 50 likely pathogenic and 50 pathogenic) were randomly selected across different genes. The evidence listed in the ClinGen Evidence Repository for these variants was independently reviewed by three experienced HKGP curators and was converted to rules following the general ACMG guideline. In these 150 variants, publications used to support the applied rules were downloaded. For rules not applied by the ClinGen expert panels, two relevant publications associated with the target variant were retrieved using the Mastermind search engine (Genomenon). These publications and any supplementary materials were processed and submitted to the LLMs as described in the supplementary methods. The results were evaluated at two levels: (i) whether each individual rule was correctly applied at the appropriate strength, and (ii) whether the overall classification of the variant matched the final classification provided by human curators. At present, human curators still need to manually aggregate the results of individual rule decisions to obtain the final classification. Development of an automated system to streamline this result aggregation process is currently underway.

### Evaluation of the optimized framework for variant reanalysis

To demonstrate the utility of our framework in variant reanalysis, 93 ClinVar variants with conflicting classifications and 57 ClinVar VUS were selected for analysis. Publications relevant to the seven literature-dependent ACMG rules were retrieved using the Mastermind search engine (Genomenon). These publications, along with their supplementary materials, were processed and submitted to the LLMs following the same workflow described above and in the supplementary methods. The reanalysis results were evaluated using the same criteria as the ClinGen-curated variants

### Statistical analysis

No statistical analysis was performed in this study.

## Data Availability

All data associated with this study are present in the paper or the Supplementary Materials. DeepSeeK-R1 and o3-mini-high are available on https://poe.com/.

## Supplementary Materials

Materials and Methods

Supplementary Figure 1. Workflow of the PVS1(RNA) prompt

Supplementary Figure 2. Workflow of the PS2/PM6 prompt

Supplementary Figure 3. Workflow of the PS3 prompt

Supplementary Figure 4. Workflow of the PS4 prompt

Supplementary Figure 5. Workflow of the PM3 prompt

Supplementary Figure 6. Workflow of the PP1 prompt

Supplementary Figure 7. Workflow of the PP4 prompt

Supplementary Table 1: Selected variants for testing PVS1(RNA)

Supplementary Table 2: Selected variants for testing PS2/PM6

Supplementary Table 3: Selected variants for testing PS3

Supplementary Table 4: Selected variants for testing PS4

Supplementary Table 5: Selected variants for testing PM3

Supplementary Table 5: Selected variants for testing PP1

Supplementary Table 6: Selected variants for testing PP4

Supplementary Table 8: Gene-disease consistency table for designing PS2/PM6 strength

Supplementary Table 9: Knowledgebase of modified ClinGen PP1 recommendation

Supplementary Table 10: Knowledgebase for PP4

Supplementary Table 11: Summary of PS4 thresholds proposed by ClinGen expert panels

Supplementary Table 12: Classification of 150 ClinGen curated variants by human curator and LLMs

Supplementary Table 13: Reclassification of 150 ClinVar variants (VUS or conflicting pathogenicity) by human curator and LLMs

## ACKNOWLEDGEMENTS

The authors would like to thank Dr Su-vui Lo, Chief Executive Officer, of the Hong Kong Genome Institute for the instrumental leadership and guidance. We also thank all patients and family members for participating in the Project. We wish to acknowledge the support of the healthcare and recruitment teams at the Partnering Centres of HKGI: The University of Hong Kong/Queen Mary Hospital, The Chinese University of Hong Kong/Prince of Wales Hospital, and Hong Kong Children’s Hospital. We also wish to thank the ACMG workgroup members and members in all ClinGen working groups and expert panels for their contribution in establishing and refining the variant classification system. Finally, the authors would like to express their gratitude to the HKGI Board of Directors and Advisory Committees and HKGP stakeholders: The Health Bureau, Hospital Authority, Department of Health and the Government of the Hong Kong SAR.

## FUNDINGS

The HKGP is a large-scale genome sequencing project funded and wholly owned by the Health Bureau of the Hong Kong Special Administrative Region (SAR) Government of China.

## AUTHOR CONTRIBUTIONS

WM, AC and BC conceptualized the project. WM, GF, JL and HW performed data collection and analysis. WM drafted the manuscript. BC supervised the study. All authors reviewed and approved the manuscript.

## COMPETING INTERESTS

The authors declare no competing interests.

## Notes

### Competing Interest Statement

The authors have declared no competing interest.

## REFERENCES AND NOTES

1. F. Liu, H. Zhou, B. Gu, X. Zou, J. Huang, J. Wu, Y. Li, S. S. Chen, Y. Hua, P. Zhou, J. Liu, C. Mao, C. You, X. Wu, Y. Zheng, L. Clifton, Z. Li, J. Luo, D. A. Clifton, Application of large language models in medicine. Nat. Rev. Bioeng. , 1–20 (2025).

2. D. Wang, S. Zhang, Large language models in medical and healthcare fields: applications, advances, and challenges. Artif. Intell. Rev. 57, 299 (2024).

3. S. Richards, N. Aziz, S. Bale, D. Bick, S. Das, J. Gastier-Foster, W. W. Grody, M. Hegde, E. Lyon, E. Spector, K. Voelkerding, H. L. Rehm, on behalf of the A. L. Q. A. Committee, Standards and guidelines for the interpretation of sequence variants: a joint consensus recommendation of the American College of Medical Genetics and Genomics and the Association for Molecular Pathology. Genet. Med. 17, 405–423 (2015).

4. D. V. Veen, C. V. Uden, L. Blankemeier, J.-B. Delbrouck, A. Aali, C. Bluethgen, A. Pareek, M. Polacin, E. P. Reis, A. Seehofnerová, N. Rohatgi, P. Hosamani, W. Collins, N. Ahuja, C. P. Langlotz, J. Hom, S. Gatidis, J. Pauly, A. S. Chaudhari, Adapted large language models can outperform medical experts in clinical text summarization. Nat. Med. 30, 1134–1142 (2024).

5. G. Xiong, Q. Jin, Z. Lu, A. Zhang, Benchmarking Retrieval-Augmented Generation for Medicine. Find. Assoc. Comput. Linguistics ACL 2024, 6233–6251 (2024).

6. D. Xu, Z. Zhang, Z. Zhu, Z. Lin, Q. Liu, X. Wu, T. Xu, X. Zhao, Y. Zheng, E. Chen, Mitigating Hallucinations of Large Language Models in Medical Information Extraction via Contrastive Decoding. Find. Assoc. Comput. Linguistics: EMNLP 2024 , 7744–7757 (2024).

7. S. Sandmann, S. Hegselmann, M. Fujarski, L. Bickmann, B. Wild, R. Eils, J. Varghese, Benchmark evaluation of DeepSeek large language models in clinical decision-making. Nat. Med. , 1–4 (2025).

8. M. Tordjman, Z. Liu, M. Yuce, V. Fauveau, Y. Mei, J. Hadjadj, I. Bolger, H. Almansour, C. Horst, A. S. Parihar, A. Geahchan, A. Meribout, N. Yatim, N. Ng, P. Robson, A. Zhou, S. Lewis, M. Huang, T. Deyer, B. Taouli, H.-C. Lee, Z. A. Fayad, X. Mei, Comparative benchmarking of the DeepSeek large language model on medical tasks and clinical reasoning. Nat. Med. , 1–6 (2025).

9. DeepSeek-AI, D. Guo, D. Yang, H. Zhang, J. Song, R. Zhang, R. Xu, Q. Zhu, S. Ma, P. Wang, X. Bi, X. Zhang, X. Yu, Y. Wu, Z. F. Wu, Z. Gou, Z. Shao, Z. Li, Z. Gao, A. Liu, B. Xue, B. Wang, B. Wu, B. Feng, C. Lu, C. Zhao, C. Deng, C. Zhang, C. Ruan, D. Dai, D. Chen, D. Ji, E. Li, F. Lin, F. Dai, F. Luo, G. Hao, G. Chen, G. Li, H. Zhang, H. Bao, H. Xu, H. Wang, H. Ding, H. Xin, H. Gao, H. Qu, H. Li, J. Guo, J. Li, J. Wang, J. Chen, J. Yuan, J. Qiu, J. Li, J. L. Cai, J. Ni, J. Liang, J. Chen, K. Dong, K. Hu, K. Gao, K. Guan, K. Huang, K. Yu, L. Wang, L. Zhang, L. Zhao, L. Wang, L. Zhang, L. Xu, L. Xia, M. Zhang, M. Zhang, M. Tang, M. Li, M. Wang, M. Li, N. Tian, P. Huang, P. Zhang, Q. Wang, Q. Chen, Q. Du, R. Ge, R. Zhang, R. Pan, R. Wang, R. J. Chen, R. L. Jin, R. Chen, S. Lu, S. Zhou, S. Chen, S. Ye, S. Wang, S. Yu, S. Zhou, S. Pan, S. S. Li, S. Zhou, S. Wu, S. Ye, T. Yun, T. Pei, T. Sun, T. Wang, W. Zeng, W. Zhao, W. Liu, W. Liang, W. Gao, W. Yu, W. Zhang, W. L. Xiao, W. An, X. Liu, X. Wang, X. Chen, X. Nie, X. Cheng, X. Liu, X. Xie, X. Liu, X. Yang, X. Li, X. Su, X. Lin, X. Q. Li, X. Jin, X. Shen, X. Chen, X. Sun, X. Wang, X. Song, X. Zhou, X. Wang, X. Shan, Y. K. Li, Y. Q. Wang, Y. X. Wei, Y. Zhang, Y. Xu, Y. Li, Y. Zhao, Y. Sun, Y. Wang, Y. Yu, Y. Zhang, Y. Shi, Y. Xiong, Y. He, Y. Piao, Y. Wang, Y. Tan, Y. Ma, Y. Liu, Y. Guo, Y. Ou, Y. Wang, Y. Gong, Y. Zou, Y. He, Y. Xiong, Y. Luo, Y. You, Y. Liu, Y. Zhou, Y. X. Zhu, Y. Xu, Y. Huang, Y. Li, Y. Zheng, Y. Zhu, Y. Ma, Y. Tang, Y. Zha, Y. Yan, Z. Z. Ren, Z. Ren, Z. Sha, Z. Fu, Z. Xu, Z. Xie, Z. Zhang, Z. Hao, Z. Ma, Z. Yan, Z. Wu, Z. Gu, Z. Zhu, Z. Liu, Z. Li, Z. Xie, Z. Song, Z. Pan, Z. Huang, Z. Xu, Z. Zhang, Z. Zhang, DeepSeek-R1: Incentivizing Reasoning Capability in LLMs via Reinforcement Learning. arXiv (2025), doi:10.48550/arxiv.2501.12948.

10. S. Li, Y. Wang, C.-M. Liu, Y. Huang, T.-W. Lam, R. Luo, AutoPM3: Enhancing Variant Interpretation via LLM-driven PM3 Evidence Extraction from Scientific Literature. *bioRxiv* , 2024.10.29.621006 (2024).

11. F. Cristofoli, M. Daja, P. E. Maltese, G. Guerri, B. Tanzi, R. Miotto, G. Bonetti, J. Miertus, P. Chiurazzi, L. Stuppia, V. Gatta, S. Cecchin, M. Bertelli, G. Marceddu, MAGI-ACMG: Algorithm for the Classification of Variants According to ACMG and ACGS Recommendations. Genes 14, 1600 (2023).

12. B. Meskó, Prompt Engineering as an Important Emerging Skill for Medical Professionals: Tutorial. J. Méd. Internet Res. 25, e50638 (2023).

13. Y. H. Ke, L. Jin, K. Elangovan, H. R. Abdullah, N. Liu, A. T. H. Sia, C. R. Soh, J. Y. M. Tung, J. C. L. Ong, C.-F. Kuo, S.-C. Wu, V. P. Kovacheva, D. S. W. Ting, Retrieval augmented generation for 10 large language models and its generalizability in assessing medical fitness. NPJ Digit. Med. 8, 187 (2025).

14. L. G. Biesecker, A. B. Byrne, S. M. Harrison, T. Pesaran, A. A. Schäffer, B. H. Shirts, S. V. Tavtigian, H. L. Rehm, C. S. V. I. W. Group, ClinGen guidance for use of the PP1/BS4 co-segregation and PP4 phenotype specificity criteria for sequence variant pathogenicity classification. Am. J. Hum. Genet. 111, 24–38 (2024).

15. S. Gilbert, J. N. Kather, A. Hogan, Augmented non-hallucinating large language models as medical information curators. NPJ Digit. Med. 7, 100 (2024).

16. V. Pejaver, A. B. Byrne, B.-J. Feng, K. A. Pagel, S. D. Mooney, R. Karchin, A. O’Donnell-Luria, S. M. Harrison, S. V. Tavtigian, M. S. Greenblatt, L. G. Biesecker, P. Radivojac, S. E. Brenner, C. S. V. I. W. Group, L. G. Biesecker, S. M. Harrison, A. A. Tayoun, J. S. Berg, S. E. Brenner, G. R. Cutting, S. Ellard, M. S. Greenblatt, P. Kang, I. Karbassi, R. Karchin, J. Mester, A. O’Donnell-Luria, T. Pesaran, S. E. Plon, H. L. Rehm, N. T. Strande, S. V. Tavtigian, S. Topper, Calibration of computational tools for missense variant pathogenicity classification and ClinGen recommendations for PP3/BP4 criteria. Am J Hum Genetics 109, 2163–2177 (2022).

17. A. Moses, P. Bhalla, A. Thompson, L. Lai, F. S. Coskun, C. M. Seroogy, M. T. de la Morena, C. A. Wysocki, N. S. C. van Oers, Comprehensive phenotypic analysis of diverse FOXN1 variants. J. Allergy Clin. Immunol. 152, 1273–1291.e15 (2023).

18. S. Firtina, F. Cipe, Y. Y. Ng, A. Kiykim, O. H. Ng, T. Sudutan, C. Aydogmus, S. Baris, G. Ozturk, E. Aydiner, A. Ozen, M. Sayitoglu, A Novel FOXN1 Variant Is Identified in Two Siblings with Nude Severe Combined Immunodeficiency. J. Clin. Immunol. 39, 144–147 (2019).

19. J. M. van de Kamp, O. T. Betsalel, S. Mercimek-Mahmutoglu, L. Abulhoul, S. Grünewald, I. Anselm, H. Azzouz, D. Bratkovic, A. de Brouwer, B. Hamel, T. Kleefstra, H. Yntema, J. Campistol, M. A. Vilaseca, D. Cheillan, M. D’Hooghe, L. Diogo, P. Garcia, C. Valongo, M. Fonseca, S. Frints, B. Wilcken, S. von der Haar, H. E. Meijers-Heijboer, F. Hofstede, D. Johnson, S. G. Kant, L. Lion-Francois, G. Pitelet, N. Longo, J. A. Maat-Kievit, J. P. Monteiro, A. Munnich, A. C. Muntau, M. C. Nassogne, H. Osaka, K. Ounap, J. M. Pinard, S. Quijano-Roy, I. Poggenburg, N. Poplawski, O. Abdul-Rahman, A. Ribes, A. Arias, J. Yaplito-Lee, A. Schulze, C. E. Schwartz, S. Schwenger, G. Soares, Y. Sznajer, V. Valayannopoulos, H. V. Esch, S. Waltz, M. M. C. Wamelink, P. J. W. Pouwels, A. Errami, M. S. van der Knaap, C. Jakobs, G. M. Mancini, G. S. Salomons, Phenotype and genotype in 101 males with X-linked creatine transporter deficiency. J. Méd. Genet. 50, 463 (2013).

20. L. M. Amendola, K. Muenzen, L. G. Biesecker, K. M. Bowling, G. M. Cooper, M. O. Dorschner, C. Driscoll, A. K. M. Foreman, K. Golden-Grant, J. M. Greally, L. Hindorff, D. Kanavy, V. Jobanputra, J. J. Johnston, E. E. Kenny, S. McNulty, P. Murali, J. Ou, B. C. Powell, H. L. Rehm, B. Rolf, T. S. Roman, J. V. Ziffle, S. Guha, A. Abhyankar, D. Crosslin, E. Venner, B. Yuan, H. Zouk, on behalf of the C. S. and D. Y. working group, G. P. Jarvik, Variant Classification Concordance using the ACMG-AMP Variant Interpretation Guidelines across Nine Genomic Implementation Research Studies. Am. J. Hum. Genet. 107, 932–941 (2020).

21. S. Althari, L. A. Najmi, A. J. Bennett, I. Aukrust, J. K. Rundle, K. Colclough, J. Molnes, A. Kaci, S. Nawaz, T. van der Lugt, N. Hassanali, A. Mahajan, A. Molven, S. Ellard, M. I. McCarthy, L. Bjørkhaug, P. R. Njølstad, A. L. Gloyn, Unsupervised Clustering of Missense Variants in HNF1A Using Multidimensional Functional Data Aids Clinical Interpretation. Am. J. Hum. Genet. 107, 670–682 (2020).

22. D. Smedley, K. R. Smith, A. R. Martin, E. A. Thomas, E. M. McDonagh, V. Cipriani, J. M. Ellingford, G. Arno, A. Tucci, J. Vandrovcova, G. Chan, H. J. Williams, T. Ratnaike, W. Wei, K. Stirrups, K. Ibanez, L. Moutsianas, M. Wielscher, A. Need, M. R. Barnes, L. Vestito, J. Buchanan, S. Wordsworth, S. Ashford, K. Rehmstrom, E. Li, G. Fuller, P. Twiss, O. Spasic-Boskovic, S. Halsall, R. A. Floto, K. Poole, A. Wagner, S. G. Mehta, M. Gurnell, N. Burrows, R. James, C. Penkett, E. Dewhurst, S. Gräf, R. Mapeta, M. Kasanicki, A. Haworth, H. Savage, DipRCPath, M. Babcock, M. G. Reese, M. Bale, E. Baple, C. Boustred, H. Brittain, A. de Burca, M. Bleda, A. Devereau, D. Halai, E. Haraldsdottir, Z. Hyder, D. Kasperaviciute, C. Patch, D. Polychronopoulos, A. Matchan, R. Sultana, M. Ryten, A. L. T. Tavares, C. Tregidgo, C. Turnbull, M. Welland, S. Wood, C. Snow, E. Williams, S. Leigh, R. E. Foulger, L. C. Daugherty, O. Niblock, I. U. S. Leong, C. F. Wright, J. Davies, C. Crichton, J. Welch, K. Woods, L. Abulhoul, P. Aurora, D. Bockenhauer, A. Broomfield, M. A. Cleary, T. Lam, M. Dattani, E. Footitt, V. Ganesan, S. Grunewald, S. Compeyrot-Lacassagne, F. Muntoni, C. Pilkington, R. Quinlivan, N. Thapar, C. Wallis, L. R. Wedderburn, A. Worth, T. Bueser, C. Compton, C. Deshpande, H. Fassihi, E. Haque, L. Izatt, D. Josifova, S. Mohammed, L. Robert, S. Rose, D. Ruddy, R. Sarkany, G. Say, A. C. Shaw, A. Wolejko, B. Habib, G. Burns, S. Hunter, R. J. Grocock, S. J. Humphray, P. N. Robinson, M. Haendel, M. A. Simpson, S. Banka, J. Clayton-Smith, S. Douzgou, G. Hall, H. B. Thomas, R. T. O’Keefe, M. Michaelides, A. T. Moore, S. Malka, N. Pontikos, A. C. Browning, V. Straub, G. S. Gorman, R. Horvath, R. Quinton, A. M. Schaefer, P. Yu-Wai-Man, D. M. Turnbull, R. McFarland, R. W. Taylor, Oc. Emer, Y. Janice, N. Katrina, H. R. Morris, J. Polke, N. W. Wood, C. Campbell, C. Camps, K. Gibson, N. Koelling, T. Lester, A. H. Németh, C. Palles, S. Patel, N. B. Roy, A. Sen, J. Taylor, P. Cacheiro, J. O. Jacobsen, E. G. Seaby, V. Davison, L. Chitty, A. Douglas, K. Naresh, D. McMullan, S. Ellard, I. K. Temple, A. D. Mumford, G. Wilson, P. Beales, M. Bitner-Glindzicz, G. Black, J. R. Bradley, P. Brennan, J. Burn, P. F. Chinnery, P. Elliott, F. Flinter, H. Houlden, M. Irving, W. Newman, S. Rahman, J. A. Sayer, J. C. Taylor, A. R. Webster, A. O. Wilkie, W. H. Ouwehand, F. L. Raymond, N. Bioresource, J. Chisholm, S. Hill, D. Bentley, R. H. Scott, T. Fowler, A. Rendon, M. Caulfield, 100,000 Genomes Pilot on Rare-Disease Diagnosis in Health Care — Preliminary Report. N. Engl. J. Med. 385, 1868–1880 (2021).

23. W. K. J. Lam, C. S. Lau, H. M. Luk, L. W. C. Au, G. C. P. Chan, W. Y. H. Chan, S. S. W. Cheng, T. H. T. Cheng, L. L. Cheung, Y. F. Cheung, J. S. C. Chong, A. T. W. Chu, C. C. Y. Chung, K. L. Chung, C. W. Fung, E. L. W. Fung, Y. Gao, S. Ho, S. P. Y. Hue, C.-H. Lee, T. L. Lee, P. H. Li, H. M. Lo, I. F. M. Lo, H. H. F. Loong, B. M. Ma, W. Ma, S. Y. Y. Pang, W.-K. Seto, S. W. K. Siu, H. So, Y. H. Tam, W. Tang, R. M. S. Wong, D. Y. H. Yap, M. L. Y. Yau, B. H. Y. Chung, S.-V. Lo, H. K. G. Project, The implementation of genome sequencing in rare genetic diseases diagnosis: a pilot study from the Hong Kong genome project. Lancet Reg. Heal.: West. Pac. 55, 101473 (2025).

24. A. Jolly, H. Du, C. Borel, N. Chen, S. Zhao, C. M. Grochowski, R. Duan, J. M. Fatih, M. Dawood, S. Salvi, S. N. Jhangiani, D. M. Muzny, A. Koch, K. Rouskas, S. Glentis, E. Deligeoroglou, F. Bacopoulou, C. A. Wise, J. E. Dietrich, I. B. V. den Veyver, A. S. Dimas, S. Brucker, V. R. Sutton, R. A. Gibbs, S. E. Antonarakis, N. Wu, Z. H. Coban-Akdemir, L. Zhu, J. E. Posey, J. R. Lupski, Rare variant enrichment analysis supports GREB1L as a contributory driver gene in the etiology of Mayer-Rokitansky-Küster-Hauser syndrome. Hum. Genet. Genom. Adv. 4, 100188 (2023).

25. C. F. Wright, P. Campbell, R. Y. Eberhardt, S. Aitken, D. Perrett, S. Brent, P. Danecek, E. J. Gardner, V. K. Chundru, S. J. Lindsay, K. Andrews, J. Hampstead, J. Kaplanis, K. E. Samocha, A. Middleton, J. Foreman, R. J. Hobson, M. J. Parker, H. C. Martin, D. R. FitzPatrick, M. E. Hurles, H. V. Firth, D. Study, Genomic Diagnosis of Rare Pediatric Disease in the United Kingdom and Ireland. N. Engl. J. Med. 388, 1559–1571 (2023).

26. M. M. Clark, A. Hildreth, S. Batalov, Y. Ding, S. Chowdhury, K. Watkins, K. Ellsworth, B. Camp, C. I. Kint, C. Yacoubian, L. Farnaes, M. N. Bainbridge, C. Beebe, J. J. A. Braun, M. Bray, J. Carroll, J. A. Cakici, S. A. Caylor, C. Clarke, M. P. Creed, J. Friedman, A. Frith, R. Gain, M. Gaughran, S. George, S. Gilmer, J. Gleeson, J. Gore, H. Grunenwald, R. L. Hovey, M. L. Janes, K. Lin, P. D. McDonagh, K. McBride, P. Mulrooney, S. Nahas, D. Oh, A. Oriol, L. Puckett, Z. Rady, M. G. Reese, J. Ryu, L. Salz, E. Sanford, L. Stewart, N. Sweeney, M. Tokita, L. V. D. Kraan, S. White, K. Wigby, B. Williams, T. Wong, M. S. Wright, C. Yamada, P. Schols, J. Reynders, K. Hall, D. Dimmock, N. Veeraraghavan, T. Defay, S. F. Kingsmore, Diagnosis of genetic diseases in seriously ill children by rapid whole-genome sequencing and automated phenotyping and interpretation. Sci. Transl. Med. 11 (2019), doi:10.1126/scitranslmed.aat6177.

27. H. Liang, B. Y. Tsui, H. Ni, C. C. S. Valentim, S. L. Baxter, G. Liu, W. Cai, D. S. Kermany, X. Sun, J. Chen, L. He, J. Zhu, P. Tian, H. Shao, L. Zheng, R. Hou, S. Hewett, G. Li, P. Liang, X. Zang, Z. Zhang, L. Pan, H. Cai, R. Ling, S. Li, Y. Cui, S. Tang, H. Ye, X. Huang, W. He, W. Liang, Q. Zhang, J. Jiang, W. Yu, J. Gao, W. Ou, Y. Deng, Q. Hou, B. Wang, C. Yao, Y. Liang, S. Zhang, Y. Duan, R. Zhang, S. Gibson, C. L. Zhang, O. Li, E. D. Zhang, G. Karin, N. Nguyen, X. Wu, C. Wen, J. Xu, W. Xu, B. Wang, W. Wang, J. Li, B. Pizzato, C. Bao, D. Xiang, W. He, S. He, Y. Zhou, W. Haw, M. Goldbaum, A. Tremoulet, C.-N. Hsu, H. Carter, L. Zhu, K. Zhang, H. Xia, Evaluation and accurate diagnoses of pediatric diseases using artificial intelligence. Nat. Med. 25, 433–438 (2019).

28. S. F. Kingsmore, R. Nofsinger, K. Ellsworth, Rapid genomic sequencing for genetic disease diagnosis and therapy in intensive care units: a review. NPJ Genom. Med. 9, 17 (2024).

29. T. Chen, C. Fan, Y. Huang, J. Feng, Y. Zhang, J. Miao, X. Wang, Y. Li, C. Huang, W. Jin, C. Tang, L. Feng, Y. Yin, B. Zhu, M. Sun, X. Liu, J. Xiang, M. Tan, L. Jia, L. Chen, H. Huang, H. Peng, X. Sun, X. Gu, Z. Peng, B. Zhu, H. Zou, L. Han, Genomic Sequencing as a First-Tier Screening Test and Outcomes of Newborn Screening. *JAMA Netw*. Open 6, e2331162 (2023).

30. S. Best, Z. Fehlberg, C. Richards, M. C. J. Quinn, S. Lunke, A. B. Spurdle, K. S. Kassahn, C. Patel, D. F. Vears, I. Goranitis, F. Lynch, A. Robertson, E. Tudini, J. Christodoulou, H. Scott, J. McGaughran, Z. Stark, Reanalysis of genomic data in rare disease: current practice and attitudes among Australian clinical and laboratory genetics services. Eur. J. Hum. Genet. 32, 1428–1435 (2024).

31. H. Zouk, W. Yu, A. Oza, M. Hawley, P. K. V. Kumar, C. Koch, L. M. Mahanta, J. B. Harley, G. P. Jarvik, E. W. Karlson, K. A. Leppig, M. F. Myers, C. A. Prows, M. S. Williams, S. T. Weiss, M. S. Lebo, H. L. Rehm, Reanalysis of eMERGE phase III sequence variants in 10,500 participants and infrastructure to support the automated return of knowledge updates. Genet. Med. 24, 454–462 (2022).

32. Z. Fehlberg, Z. Stark, S. Best, Reanalysis of genomic data, how do we do it now and what if we automate it? A qualitative study. Eur. J. Hum. Genet. 32, 521–528 (2024).

33. V. Cipriani, L. Vestito, E. F. Magavern, J. O. B. Jacobsen, G. Arno, E. R. Behr, K. A. Benson, M. Bertoli, D. Bockenhauer, M. R. Bowl, K. Burley, L. F. Chan, P. Chinnery, P. J. Conlon, M. A. Costa, A. E. Davidson, S. J. Dawson, E. A. E. Elhassan, S. E. Flanagan, M. Futema, D. P. Gale, S. García-Ruiz, C. G. Corcia, H. R. Griffin, S. Hambleton, A. R. Hicks, H. Houlden, R. S. Houlston, S. A. Howles, R. Kleta, I. Lekkerkerker, S. Lin, P. Liskova, H. H. Mitchison, H. Morsy, A. D. Mumford, W. G. Newman, R. Neatu, E. A. O’Toole, A. C. M. Ong, A. T. Pagnamenta, S. Rahman, N. Rajan, P. N. Robinson, M. Ryten, O. Sadeghi-Alavijeh, J. A. Sayer, C. L. Shovlin, J. C. Taylor, O. Teltsh, I. Tomlinson, A. Tucci, C. Turnbull, A. M. van Eerde, J. S. Ware, L. M. Watts, A. R. Webster, S. K. Westbury, S. L. Zheng, M. Caulfield, D. Smedley, Rare disease gene association discovery in the 100,000 Genomes Project. Nature , 1–9 (2025).

34. E. G. Seaby, H. L. Rehm, A. O’Donnell-Luria, Strategies to Uplift Novel Mendelian Gene Discovery for Improved Clinical Outcomes. Front. Genet. 12, 674295 (2021).

35. K. D. Kernohan, K. M. Boycott, The expanding diagnostic toolbox for rare genetic diseases. Nat. Rev. Genet. 25, 401–415 (2024).

36. P. Dai, A. Honda, L. Ewans, J. McGaughran, L. Burnett, M. Law, T. G. Phan, Recommendations for next generation sequencing data reanalysis of unsolved cases with suspected Mendelian disorders: A systematic review and meta-analysis. Genet. Med. 24, 1618–1629 (2022).

37. ACGS, ACGS best practice guidelines for variant classification in rare disease. (2024) (available at https://www.acgs.uk.com/media/12533/uk-practice-guidelines-for-variant-classification-v12-2024.pdf).

38. G. Houge, A. Laner, S. Cirak, N. de Leeuw, H. Scheffer, J. T. den Dunnen, Stepwise ABC system for classification of any type of genetic variant. Eur. J. Hum. Genet. 30, 150– 159 (2022).

39. R. J. Schmidt, M. Steeves, P. Bayrak-Toydemir, K. A. Benson, B. P. Coe, L. K. Conlin, M. Ganapathi, J. Garcia, M. H. Gollob, V. Jobanputra, M. Luo, D. Ma, G. Maston, K. McGoldrick, T. B. Palculict, T. Pesaran, T. I. Pollin, E. Qian, H. L. Rehm, E. R. Riggs, S. L. P. Schilit, P. I. Sergouniotis, T. Tvrdik, N. Watkins, L. Zec, W. Zhang, M. S. Lebo, C. L. P. A. W. Group, A. Byrne, A. Spurdle, B. Palculict, B. Coe, M. Deqiong, E. Lyon, E. Groopman, E. Qian, E. Puffenberger, E. Riggs, F. Couch, G. Maston, H. Dziadzio, J. Harraway, J. Mester, J. Garcia, J. Lerner-Ellis, K. Benson, K. Avello, K. McGoldrick, L. Conlin, L. Zec, M. Steeves, M. Richardson, M. Lebo, M. Kelly, M. Gollob, M. Luo, M. Ganapathi, N. Watkins, N. Niu, P. Sergouniotis, P. Bayrak-Toydemir, R. Schmidt, S. Schilit, S. Richards, T. Pesaran, T. Pollin, V. Jobanputra, W. Zhang, W. Chen, Y. Fan, Recommendations for risk allele evidence curation, classification, and reporting from the ClinGen Low Penetrance/Risk Allele Working Group. Genet. Med. 26, 101036 (2024).

40. E. R. Riggs, E. F. Andersen, A. M. Cherry, S. Kantarci, H. Kearney, A. Patel, G. Raca, D. I. Ritter, S. T. South, E. C. Thorland, D. Pineda-Alvarez, S. Aradhya, C. L. Martin, Technical standards for the interpretation and reporting of constitutional copy-number variants: a joint consensus recommendation of the American College of Medical Genetics and Genomics (ACMG) and the Clinical Genome Resource (ClinGen). Genet Med 22, 245–257 (2020).

41. P. Horak, M. Griffith, A. M. Danos, B. A. Pitel, S. Madhavan, X. Liu, C. Chow, H. Williams, L. Carmody, L. Barrow-Laing, D. Rieke, S. Kreutzfeldt, A. Stenzinger, D. Tamborero, M. Benary, P. S. Rajagopal, C. M. Ida, H. Lesmana, L. Satgunaseelan, J. D. Merker, M. Y. Tolstorukov, P. V. Campregher, J. L. Warner, S. Rao, M. Natesan, H. Shen, J. Venstrom, S. Roy, K. Tao, R. Kanagal-Shamanna, X. Xu, D. I. Ritter, K. Pagel, K. Krysiak, A. Dubuc, Y. M. Akkari, X. S. Li, J. Lee, I. King, G. Raca, A. H. Wagner, M. M. Li, S. E. Plon, S. Kulkarni, O. L. Griffith, D. Chakravarty, D. Sonkin, Standards for the classification of pathogenicity of somatic variants in cancer (oncogenicity): Joint recommendations of Clinical Genome Resource (ClinGen), Cancer Genomics Consortium (CGC), and Variant Interpretation for Cancer Consortium (VICC). Genet. Med. 24, 986–998 (2022).

42. P. Hager, F. Jungmann, R. Holland, K. Bhagat, I. Hubrecht, M. Knauer, J. Vielhauer, M. Makowski, R. Braren, G. Kaissis, D. Rueckert, Evaluation and mitigation of the limitations of large language models in clinical decision-making. Nat. Med. 30, 2613–2622 (2024).

43. B. Zhao, Y. Zong, L. Zhang, T. Hospedales, Benchmarking Multi-Image Understanding in Vision and Language Models: Perception, Knowledge, Reasoning, and Multi-Hop Reasoning. arXiv (2024), doi:10.48550/arxiv.2406.12742.

44. H. Wei, C. Liu, J. Chen, J. Wang, L. Kong, Y. Xu, Z. Ge, L. Zhao, J. Sun, Y. Peng, C. Han, X. Zhang, General OCR Theory: Towards OCR-2.0 via a Unified End-to-end Model. arXiv (2024), doi:10.48550/arxiv.2409.01704.

45. W. Zhao, H. Feng, Q. Liu, J. Tang, S. Wei, B. Wu, L. Liao, Y. Ye, H. Liu, W. Zhou, H. Li, C. Huang, TabPedia: Towards Comprehensive Visual Table Understanding with Concept Synergy. arXiv (2024), doi:10.48550/arxiv.2406.01326.

46. J. Yang, C. Liu, W. Deng, D. Wu, C. Weng, Y. Zhou, K. Wang, Enhancing phenotype recognition in clinical notes using large language models: PhenoBCBERT and PhenoGPT. Patterns 5, 100887 (2023).

47. X. Yang, A. Chen, N. PourNejatian, H. C. Shin, K. E. Smith, C. Parisien, C. Compas, C. Martin, A. B. Costa, M. G. Flores, Y. Zhang, T. Magoc, C. A. Harle, G. Lipori, D. A. Mitchell, W. R. Hogan, E. A. Shenkman, J. Bian, Y. Wu, A large language model for electronic health records. *npj Digit*. Med. 5, 194 (2022).

48. L. C. Walker, M. de la Hoya, G. A. R. Wiggins, A. Lindy, L. M. Vincent, M. T. Parsons, D. M. Canson, D. Bis-Brewer, A. Cass, A. Tchourbanov, H. Zimmermann, A. B. Byrne, T. Pesaran, R. Karam, S. M. Harrison, A. B. Spurdle, C. S. V. I. W. Group, L. G. Biesecker, S. M. Harrison, A. A. Tayoun, J. S. Berg, S. E. Brenner, G. R. Cutting, S. Ellard, M. S. Greenblatt, P. Kang, I. Karbassi, R. Karchin, J. Mester, A. O’Donnell-Luria, T. Pesaran, S. E. Plon, H. L. Rehm, N. T. Strande, S. V. Tavtigian, S. Topper, Using the ACMG/AMP framework to capture evidence related to predicted and observed impact on splicing: Recommendations from the ClinGen SVI Splicing Subgroup. Am. J. Hum. Genet. 110, 1046– 1067 (2023).

49. A. N. A. Tayoun, T. Pesaran, M. T. DiStefano, A. Oza, H. L. Rehm, L. G. Biesecker, S. M. Harrison, C. S. V. I. W. G. (ClinGen SVI), Recommendations for interpreting the loss of function PVS1 ACMG/AMP variant criterion. Hum Mutat 39, 1517–1524 (2018).

50. J. Xiang, J. Peng, S. Baxter, Z. Peng, AutoPVS1: An automatic classification tool for PVS1 interpretation of null variants. Hum. Mutat. 41, 1488–1498 (2020).

51. ClinGen Sequence Variant Interpretation Recommendation for de novo Criteria (PS2/PM6) -Version 1.1. Date Approved: March 18, 2018, updated May 5, 2021.

52. L. G. Biesecker, A. B. Byrne, S. M. Harrison, T. Pesaran, A. A. Schäffer, B. H. Shirts, S. V. Tavtigian, H. L. Rehm, C. S. V. I. W. Group, ClinGen guidance for use of the PP1/BS4 co-segregation and PP4 phenotype specificity criteria for sequence variant pathogenicity classification. Am. J. Hum. Genet. 111, 24–38 (2024).

